# Cost Effectiveness of Newborn Screening for Spinal Muscular Atrophy in England

**DOI:** 10.1101/2023.02.09.23285715

**Authors:** Diana Weidlich, Laurent Servais, Imran Kausar, Ruth Howells, Matthias Bischof

## Abstract

**Introduction:** We sought to evaluate the cost effectiveness of newborn screening (NBS) versus no NBS for 5q spinal muscular atrophy (SMA) in England.

**Methods:** A cost-utility analysis using a combination of decision tree and Markov model structures was developed to estimate the lifetime health effects and costs of NBS for SMA, compared with no NBS, from the perspective of the National Health Service (NHS) in England. A decision tree was designed to capture NBS outcomes, and Markov modelling was used to project long-term health outcomes and costs for each patient group following diagnosis. Model inputs were based on existing literature, local data, and expert opinion. Sensitivity and scenario analyses were conducted to assess the robustness of the model and the validity of the results.

**Results:** The introduction of NBS for SMA in England is estimated to identify approximately 56 (96% of cases) infants with SMA per year. Base-case results indicate that NBS is dominant (less costly and more effective) than a scenario without NBS, with a yearly cohort of newborns accruing incremental savings of £62,191,531 and an estimated gain in quality-adjusted life-years of 529 years over their lifetime. Deterministic and probabilistic sensitivity analyses demonstrated the robustness of the base-case results.

**Conclusions:** NBS improves health outcomes for patients with SMA and is less costly compared with no screening; therefore, it is a cost-effective use of resources from the perspective of the NHS in England.

## INTRODUCTION

Spinal muscular atrophy (SMA) is a rare genetic disorder characterized by progressive muscle weakness and atrophy, respiratory failure, and in the most severe cases, death in children younger than 2 years of age [1, 2].

More than 90% of SMA is caused by the loss of the *survival motor neuron 1 (SMN1)* gene from chromosome 5 (5q13) which leads to irreversible degeneration of motor neurons [1]. 5q SMA, herein further referred to as SMA, is classified into four types based on age of onset and maximum motor milestones achieved. SMA type 1 is the most common (accounting for 50–60% of cases) [3, 4] and most severe form of SMA, with onset in early infancy. SMA type 1 is characterized by a rapid decline of motor and respiratory function, typically leading to death or permanent assisted ventilation (PAV) before 2 years of age if left untreated [1, 2, 5]. SMA types 2 and 3 are characterized by stalled gross motor development, which causes a spectrum of symptoms such as an inability to stand or walk (type 2), or ambulation loss later in life (type 3) [1, 2]. SMA type 4 represents just <5% of SMA cases and is the least severe form of the disease, with patients retaining ambulation but with proximal weakness of arms and legs later in life [1]. The severity of the disease is mostly driven by the number of copies of *SMN2*, a nearly identical gene to *SMN1,* from which only a limited amount of SMN protein is produced.

The incidence of SMA is 1 in 10,000 live births [6–9], suggesting that approximately 62 infants are born with SMA per year in England. It is estimated that between 668 and 1,336 children and adults are living with SMA in the UK, with a worldwide prevalence ratio of 1 to 2 people per 100,000 [10].

Novel targeted treatments for SMA can prevent loss of motor neurons soon after birth, thereby preventing disease progression. In the United Kingdom, three disease-modifying treatments (DMTs), onasemnogene abeparvovec, nusinersen, and risdiplam, have been approved and are reimbursed for the treatment of SMA. These DMTs demonstrate promise when administered early, ideally prior to symptom onset, to achieve as close to a functional cure as possible [11–20]. More motor neurons are irreversibly lost with later treatment imitation [1]. Patients with SMA symptoms at the time of treatment will likely require respiratory, nutritional, or musculoskeletal support to maximize functional abilities [13, 17, 18, 21–23].

Early diagnosis of SMA through newborn screening (NBS) enables prompt treatment initiation and is critical for optimizing clinical outcomes for infants with SMA [23–26]. Although some infants identified by NBS are already symptomatic at diagnosis [24, 26, 27], implementing NBS would help all infants at risk for SMA to be identified and treated early, avoiding delays in treatment and irreversible loss of motor neurons. Treatment of infants with SMA identified by NBS is associated with lower medical costs and societal burden than for those patients diagnosed and treated following symptom onset [28]. NBS for SMA has been introduced or is under consideration in several countries [29–34].

It is important for decision-makers to determine if NBS for SMA offers value for money to the health care system. This evaluation aimed to assess the cost effectiveness of NBS for SMA and immediate treatment with DMTs compared with a scenario without NBS and symptomatic diagnosis and treatment in England.

## METHODS

### Population cohort

A total of 585,195 newborns were included in the model based on the number of live births in England in 2020[35]. The model compared two population cohorts: NBS (patients identified with SMA who were either symptomatic or presymptomatic at the time of screening) and no NBS (patients with SMA who were symptomatic at the time of diagnosis).

### Model structure and assumptions

A cost-utility analysis using a combination of decision tree and Markov model structures (**Fig. 1**) was conducted to estimate the lifetime health effects and costs of NBS compared with no NBS. A decision tree was designed to capture NBS outcomes, and Markov modelling was used to project long-term health outcomes and costs for each patient group following diagnosis. An earlier version of the model was published by Velikanova et al [33]. The Markov model included the following six health states: within a broad range of normal development (BRND) (A state), walking (B state), sitting (C state), not sitting (D state), PAV (E state) and death. All patients were assumed to be treated in the first 6 months after diagnosis (assumed to be within 6 months, 18 months, and 4 years of age for patients with SMA types 1, 2 and 3, respectively). Infants identified by NBS at risk for SMA were assumed to receive treatment shortly after birth.

**Fig. 1.**
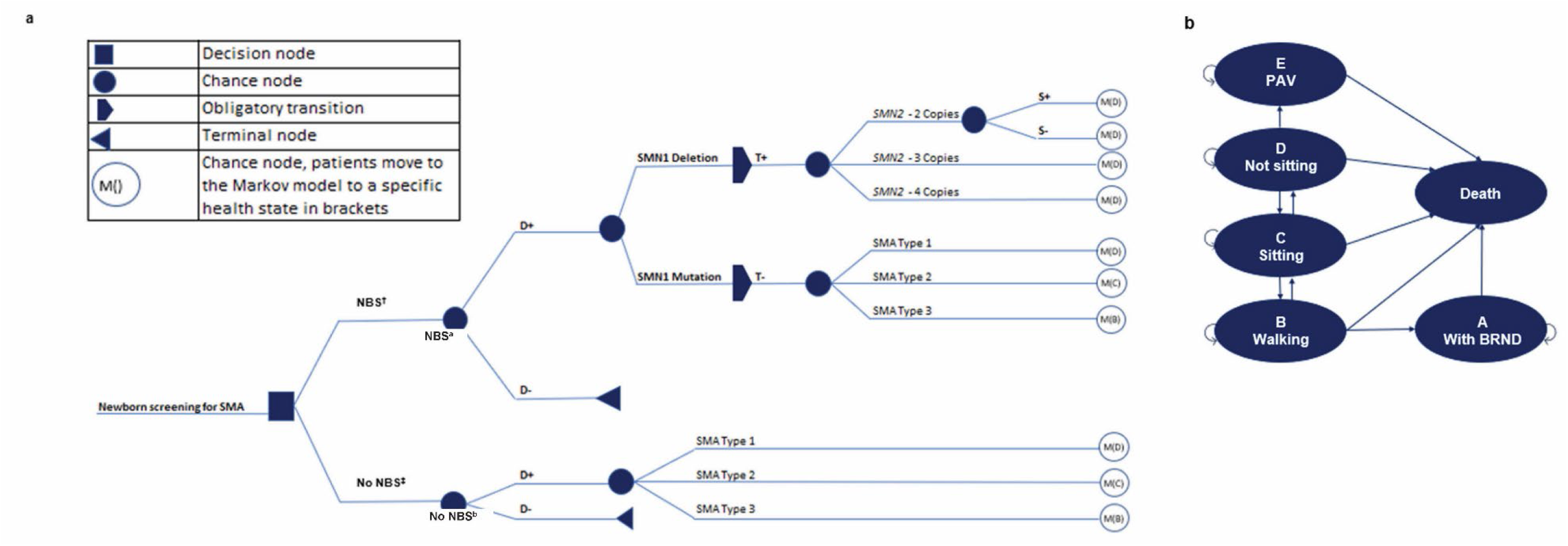
Two-part model: decision tree (a) followed by a Markov model (b) *BRND* broad range of normal development; *D+* patients with SMA; *D-* patients without SMA; *NBS* newborn screening; *PAV* permanent assisted ventilation; *S+* symptoms present; *S-* no symptoms; *SMA* spinal muscular atrophy; *SMN* survival motor neuron; *T+* positive test; *T-* negative test. ^a^Short-term model. ^b^Long-term model.

Patients in the model entered a specific Markov model health state after the decision tree, depending on diagnosis (demonstrated by M() in **Fig. 1**). All possible transitions in the Markov model are represented by arrows in **Fig. 1**. Upon achievement of motor milestones, patients were transitioned to the next health state in the next model cycle, and it was assumed that motor milestone achievement would be maintained in treated patients until death. Untreated patients in the model could lose milestones, such as independent sitting or walking. Based on data from a natural history study in SMA, it was assumed that 24% of patients with SMA type 2 with the ability to sit would lose this milestone between 0.7 and 29.1 years, and 9% of patients with SMA type 3 would lose this milestone between 15.5 and 40.5 years [36]. Of patients with SMA type 3 who were able to walk, it was assumed that 51% would lose this milestone between 2.5 and 65.7 years [36]. Patients could transition to death from any health state [36]. This Markov model has also been used to model long-term outcomes for SMA type 1 in health technology assessment submissions and other publications [33, 37, 38].

A 6-month model cycle was used for the first six cycles, followed by yearly cycles to capture changes in childhood development and milestone achievement. A lifetime time horizon was modelled for the base-case analysis (from birth/treatment initiation to age 100 years), and a discount rate of 3.5% was applied for costs and outcomes.

### Model inputs

Model inputs were based on existing literature, local data, and expert opinion.

#### Patient distribution

A total cohort of 585,195 infants was included in the model based on the number of live births in England in 2020 [35]. The incidence of 5q SMA is 1 in 10,000 live births [6–9]; homozygous deletion of *SMN1* accounts for 96% of SMA cases, and 4% of cases have a point mutation in *SMN1* [39].

In the model, of infants identified by NBS at risk for SMA who were presymptomatic, it was assumed that 46.7%, 25%, and 28.3% had two, three, or four copies of *SMN2*, respectively [24, 26, 29, 40–44]. Of infants identified by NBS at risk for SMA who were symptomatic, 58%, 29%, and 13% were assumed to have SMA types 1, 2 and 3, respectively [45]. Based on expert opinion, it was assumed that 40% of patients with two copies of *SMN2* became symptomatic by the time they received treatment (before age 6 months). SMA caused by *SMN1* point mutations was assumed to be undetectable because of testing limitations [46].

#### Treatment pattern

The proportion of patients with SMA (detected before or after symptom onset) receiving treatment by SMA type and copy number is presented in **Table 1**.

**Table 1.** Model inputs: Treatment patterns.

|  | **Onasemnogene**<br>**abeparvovec** | **Nusinersen** | **Risdiplam** | **BSC** |
| --- | --- | --- | --- | --- |
| **Presymptomatically detected<sup>a</sup> (%)** |  |  |  |  |
| *SMN2* two copies | 93 | 6 | 0 | 1 |
| *SMN2* three copies | 93 | 6 | 0 | 1 |
| *SMN2* four copies | 0 | 6 | 50 | 44 |
| **Symptomatically detected<sup>a</sup> (%)** |  |  |  |  |
| Type 1 | 56 | 2 | 22 | 20 |
| Type 2 | 0 | 10 | 90 | 0 |
| Type 3 | 0 | 10 | 90 | 0 |
| **Patients identified via screening but treated symptomatically<sup>a</sup>** |  |  |  |  |
| *SMN2* two copies | 93 | 6 | 0 | 1 |

*BSC* best supportive care; *SMN2* survival motor neuron 2 gene.
<sup>a</sup>Based on expert opinion.

#### Clinical inputs

Short-term efficacy data from relevant clinical trials provided milestone achievements for presymptomatically and symptomatically detected patients, as well as patients identified by NBS but who received treatment following symptom onset, for the first 3 years of the Markov model (**Table 2**). Because of a lack of available data for presymptomatic infants with four copies of *SMN2*, efficacy data for patients with three *SMN2* copies were applied. For patients identified by NBS but who received treatment following symptom onset, the clinical trajectory of an SMA type 3 patient was used. These assumptions were based on clinical input. Long-term survival data for each health state were extrapolated from existing literature (**Table 3**).

**Table 2.**
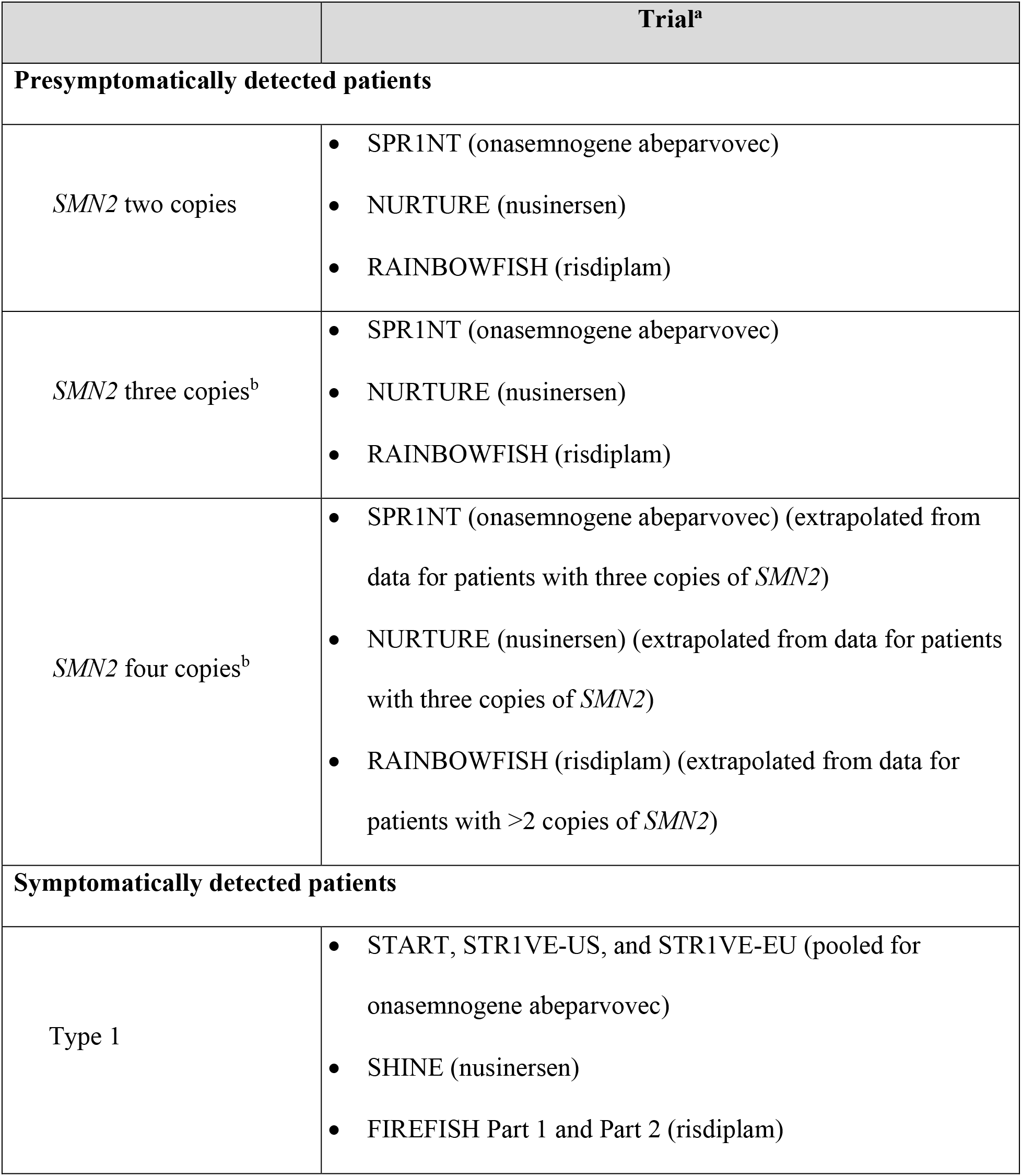
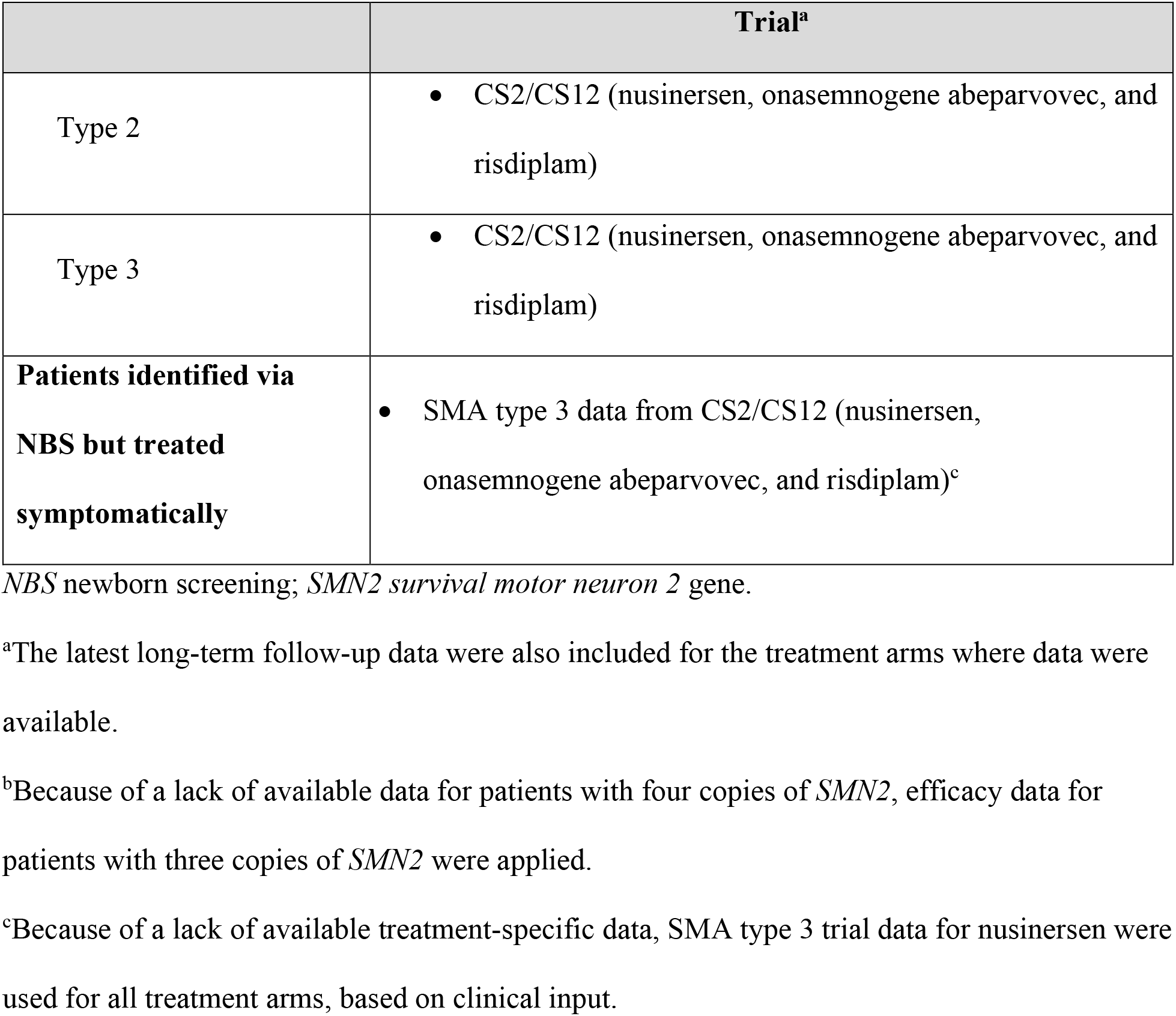
Model inputs: Short-term efficacy data used for the first 3 years of the Markov model

**Table 3.** Model inputs: Long-term efficacy data

| Health state | Description | Source<sup>a</sup> |
| --- | --- | --- |
| E state | PAV (non-invasive only) | Gregoretti et al., 2013 [47] |
| D state | Not sitting | Kolb SJ, et al., 2017 [48] |
| C state | Sitting | Zerres et al., 1997 [49] |
| B state | Walking | General population life expectancy in England, ONS National Life Tables for England 2018–2020 [50] |
| A state | Walking and within BRND | General population's life expectancy in England, ONS National Life Tables for England 2018–2020 [50] |

*BRND* broad range of normal development; *ONS* Office for National Statistics; *PAV* permanent-assisted ventilation.
<sup>a</sup>For the sitting health state, Wijngaarde et al. (2020) was used for sensitivity analysis [55].

#### Resource use, cost, and utilities

The cost of each heel prick screening test was £4.54 (a Dutch value, which is in line with other sources in Europe, converted to GBP because of lack of UK-specific data) [51]. The confirmatory genetic test was assumed to be £1,200 (based on prices from Oxford Genetic Laboratories, assuming both gene sequencing and multiplex ligation-dependent probe amplification are needed [based on the test for Duchenne/Becker muscular dystrophy]) [52].

Treatment and administration costs were based on the UK list prices and the latest National Health Service (NHS) reference costs (2019/2020) [53]. SMA care costs were based on a UK health care resource utilization (HCRU) study updated with 2019/2020 costs [37]. All costs were presented in 2021/2022 GBP values (where required, costs were inflated to 2021 values using Personal Social Services Research Unit’s NHS Cost Inflation Index [54]). Utilities were based on published literature and clinical expert input. These were the preferred values used by the National Institute for Health and Care Excellence (NICE) Evidence Review Group in their appraisal of onasemnogene abeparvovec in the United Kingdom [37] and by the Institute for Clinical and Economic Review in the United States (**Table 4**).

**Table 4.**
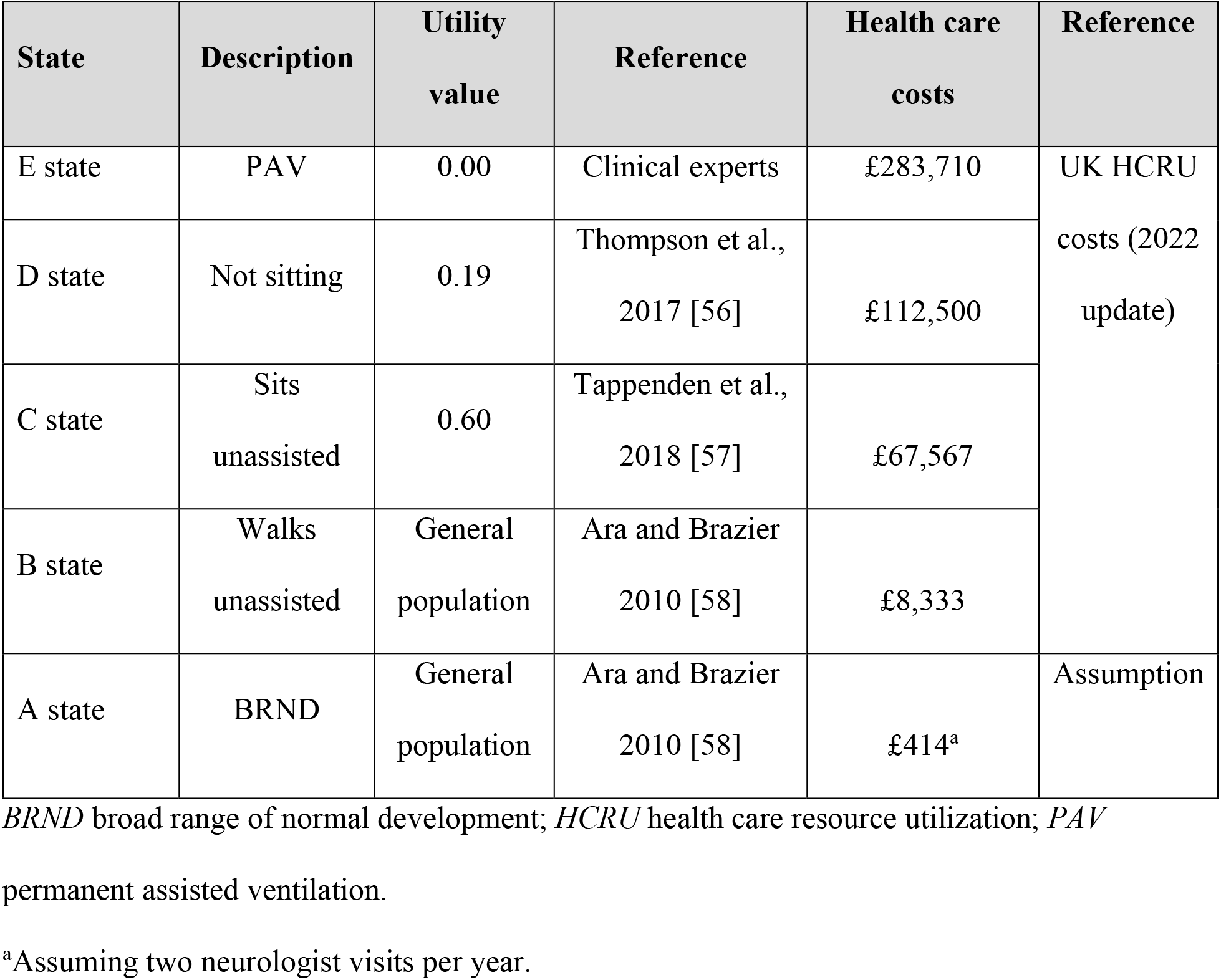
Model inputs: health care costs and utilities

### Sensitivity analyses

To assess the robustness of the model and parameters, sensitivity analyses were completed. A deterministic (univariate) sensitivity analysis (DSA) was conducted to evaluate the impact of parameter uncertainty by varying parameter values and reporting the effect on the cost-effectiveness outcomes. The probabilistic sensitivity analysis (PSA) was performed (with 1,000 iterations) to assess parametric uncertainty.

### Scenario analyses

The scenario analyses tested key model assumptions and provided an analysis of how robust the base-case incremental cost-effectiveness ratio (ICER) was to key parameters in the model.

The discount rate, time horizon, and analysis perspective (payer and societal) used in the model were assessed in the scenario analyses. Indirect, caregiver, and transportation costs were applied in the societal perspective scenario. Inputs (costs) and assumptions applied are presented in the supplementary material. An additional scenario looked at the impact of informing the survival of C state patients using a more recent natural history study of survival in patients with SMA [55], although based on a smaller group of SMA patients (n=307 [55] vs. n=569 in the study that informed the base-case analysis [49]).

## RESULTS

### Newborn screening outcomes

NBS is estimated to identify approximately 56 infants per year at risk for SMA, which is 96% of all SMA patients (4% are assumed to have an *SMN1* point mutation that is assumed undetectable by NBS owing to test limitations [39]) in England. We estimate that 46 of these patients will be asymptomatic at the time of treatment, and 10 patients will be symptomatic, even if identified by NBS (**Table 5**).

**Table 5.** Newborn screening outcomes

| Outcome | NBS | No NBS |
| --- | --- | --- |
| **Number of tests performed** | 585,254 | 58.5 |
| 1st tier – heel-prick test | 585,195 | 0 |
| 2nd tier – confirmatory genetic test | 58.5 | 58.5 |
| **Number of cases treated** | 58.5 | 58.5 |
| Patients identified and treated presymptomatically | 45.8 | 0.0 |
| Patients identified before symptom onset but treated symptomatically | 10.5<sup>a</sup> | 0.0 |
| Patients identified and treated symptomatically | 2.2<sup>b</sup> | 58.5 |

NBS newborn screening; *SMN1* survival motor neuron 1 gene; *SMN2* survival motor neuron 2 gene.
<sup>a</sup>40% of SMA patients with two copies of *SMN2* are assumed to become symptomatic by the time they receive treatment.
<sup>b</sup>4% of patients with SMA are assumed to have an *SMN1* point mutation and are thus not detected by qPCR-based newborn screening.

### Base-case results

In the base-case analysis, over the lifetime of a newborn cohort identified (yearly), total costs for NBS versus no NBS were £160,068,073 and £222,259,604, respectively, with an incremental cost savings of £62,191,531 for the NBS cohort (**Table 6**). The introduction of NBS over the lifetime of a newborn cohort identified per year was associated with total quality-adjusted life-years (QALYs) of 1,140 versus 611 for no NBS, thereby providing an incremental gain of 529 QALYs. NBS was associated with 1,346 life-years (LY) (vs 924 with no NBS) and incremental LYs of 423 over the lifetime of a newborn cohort identified per year. Base-case results indicate that NBS is dominant (less costly and more effective) compared with the scenario without NBS, with an ICER of –£117,541 per QALY (**Table 6**).

**Table 6.** Base-case results (payer perspective and discounted at 3.5% p.a.)

| Strategy | Total costs (£) | Total LYs | Total QALYs | Incremental costs (£) | Incremental LYs | Incremental QALYs | ICER (£ per QALY) |
| --- | --- | --- | --- | --- | --- | --- | --- |
| **NBS** | £160,068,073 | 1,346 | 1,140 | −£62,191,531 | 423 | 529 | −117,541<br>(dominant) |
| **No NBS** | £222,259,604 | 924 | 611 |  |  |  |  |

*ICER* incremental cost-effectiveness ratio; *LY* life-year; *NBS* newborn screening; *p.a* per annum; *QALY* quality-adjusted life year.

To provide further insight on the main drivers of health gains and health care cost savings associated with NBS and early treatment, the proportion of patients in each of the six health states of the model (*see* **Fig. 1**) was assessed at different time points to follow children’s development over time. The results of this analysis under NBS and no NBS are provided in **Fig. S1** and **Fig. S2** of the supplementary material. With NBS and early treatment, approximately 80% of children with SMA will likely sit and walk independently, as opposed to approximately 20% of children in the current situation, in which no NBS is available, from the age of 5 years old onward.

This difference in motor milestone achievements will lead to a substantially longer and improved quality of life in SMA patients treated early because of NBS, as well as a drastic reduction in costly HCRU over their lifetimes (**Table S7** in supplementary material provides economic outcomes per SMA patient), and demonstrates that NBS and early treatment are expected to provide to each patient with SMA on average an additional 32 years at full health when compared with no NBS, where children are treated at symptom onset. Implementation of NBS will also drastically reduce the costs associated with hospital admissions, breathing equipment, and other costly health care services. When also considering the reduction in drug acquisition costs owing to the different treatment patterns used for treating presymptomatic versus symptomatic patients (*see* **Table 1**), NBS with early treatment is expected to generate a (discounted) cost savings, net of population-level screening costs, of more than £1,000,000 per SMA patient compared with no NBS.

**Table 7.** Mean probabilistic results (payer perspective and discounted)

| Strategy | Total costs<br>(£) | Total<br>QALYs | Incremental costs<br>(£) | Incremental<br>QALYs | ICER (£ per<br>QALY) |
| --- | --- | --- | --- | --- | --- |
| **NBS** | £162,664,662 | 1,131 | –£59,318,947 | 526 | –£112,811<br>(dominant) |
| **No NBS** | £221,983,609 | 606 |  |  |  |

*ICER* incremental cost-effectiveness ratio; *NBS* newborn screening; *QALY* quality-adjusted life year.

### Deterministic sensitivity analysis

All DSA results indicated that NBS was dominant versus no NBS (**Fig. 2**). For all parameters varied in DSA, the ICER was dominant, indicating the robustness of the base-case results. The parameters that had the largest impact on the ICER were the general population utility intercept values, C state utility value, and resource use for ventilated patients in the C state. DSA results for all the other inputs were within ±4.4% around the base-case result.

**Fig. 2.**
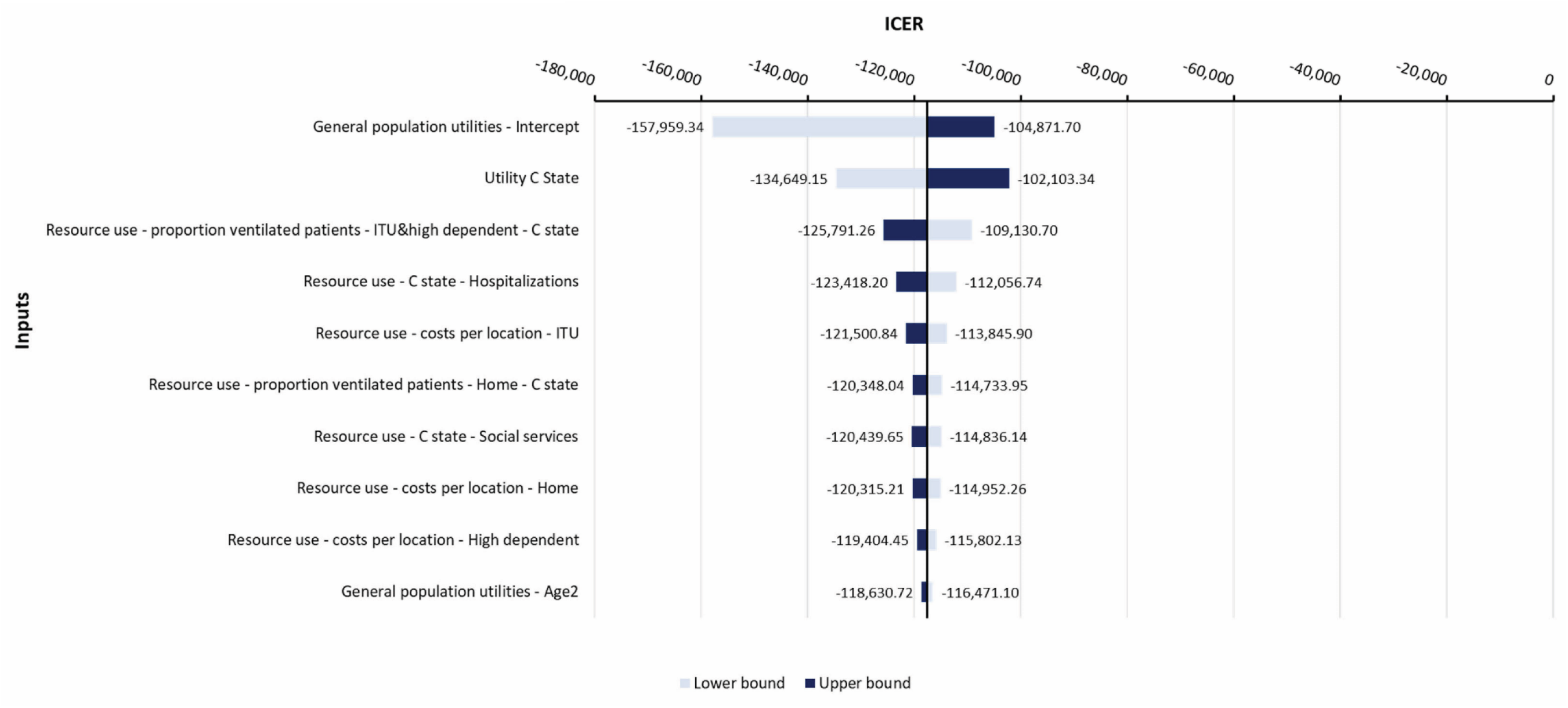
**Deterministic sensitivity analysis** *ICER* incremental cost-effectiveness ratio; *ITU* intensive care unit; *NBS* newborn screening.

### Probabilistic sensitivity analysis

Results of the PSA are presented in **Table 7** and **Fig. 3**. The PSA indicated that NBS is dominant versus no NBS, with a mean incremental cost of –£59,318,947 and a mean ICER of –£112,811 (**Table 7**), indicating the robustness of the base-case results. All simulated ICERs fall below willingness-to-pay thresholds of £20,000, £30,000, and £100,000 per QALY (**Fig. 3**) [59].

**Fig. 3.**
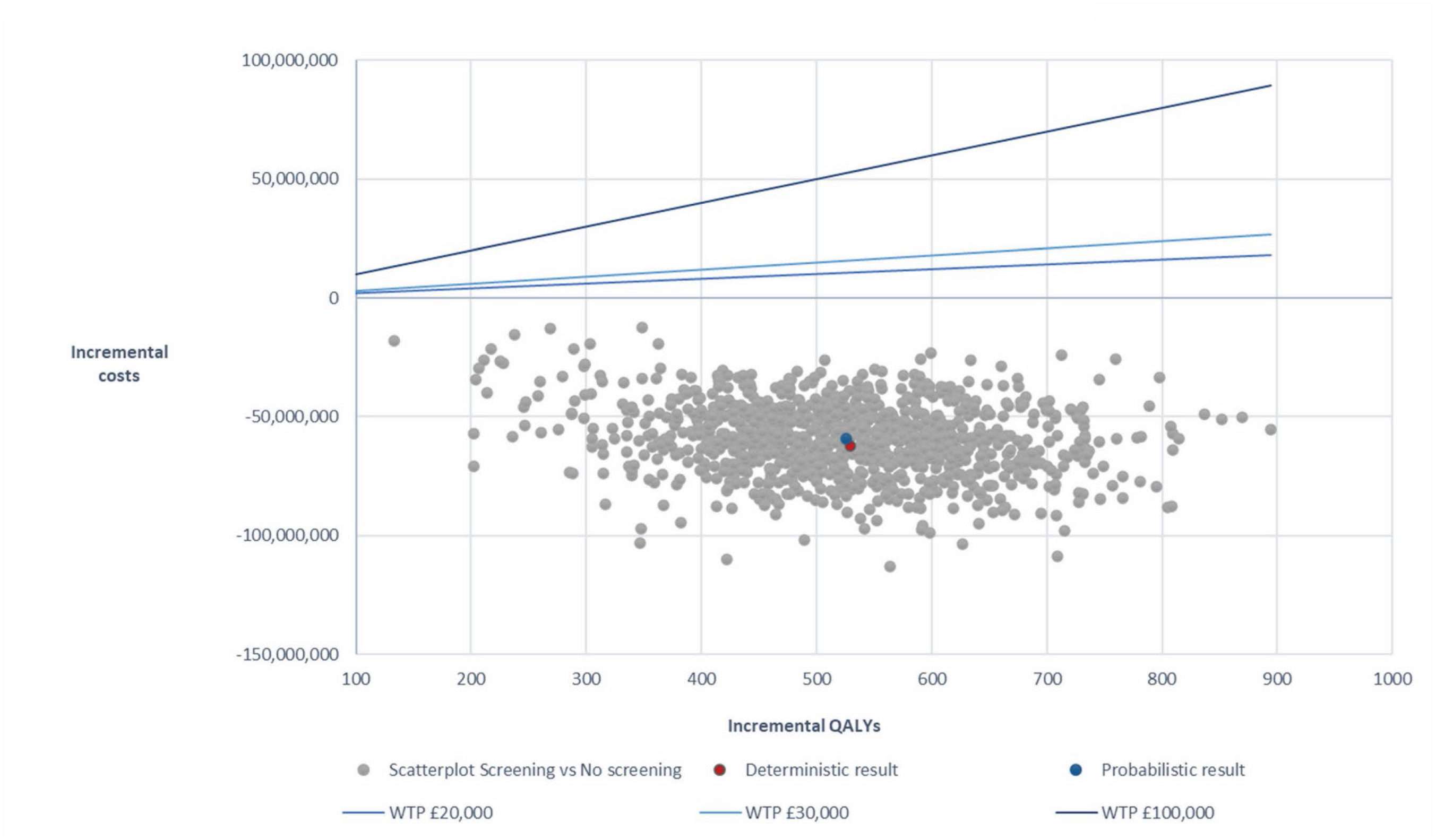
Incremental cost-effectiveness plane with willingness-to-pay thresholds *QALYs* quality-adjusted life years; *WTP* willingness-to-pay.

### Scenario analyses

Scenario analyses were performed by: (1) varying the discount rate (set to 1.5%), (2) switching lifetime time horizons (using 10 and 50 years respectively), (3) changing the data source informing the survival for patients in C state [55], and (4) incorporating societal cost in terms of lost productivity for the patients, their caregivers, and transport costs (societal perspective). All scenario analyses demonstrated that NBS was dominant compared with no NBS (**Table 8**).

**Table 8.** Scenario analysis results

| Strategy | Total costs<br>(£) | Total<br>LYs | Total<br>QALYs | Incremental costs<br>(£) | Incremental<br>LYs | Incremental<br>QALYs | ICER (£ per<br>QALY) |
| --- | --- | --- | --- | --- | --- | --- | --- |
| Discount rate of 1.5% |  |  |  |  |  |  |  |
| NBS | £211,956,043 | 2,265 | 1,924 | −£98,065,394 | 920 | 1,006 | −£97,485<br>(dominant) |
| No NBS | £310,021,437 | 1,345 | 918 |  |  |  |  |
| Lifetime time horizon of 10 years |  |  |  |  |  |  |  |
| NBS | £108,267,465 | 449 | 339 | −£4,778,101 | 49 | 105 | −£45,567<br>(dominant) |
| No NBS | £113,045,566 | 401 | 234 |  |  |  |  |
| Lifetime time horizon of 50 years |  |  |  |  |  |  |  |
| NBS | £151,618,768 | 1,192 | 1,016 | −£61,117,353 | 314 | 440 | −£139,009<br>(dominant) |
| No NBS | £212,736,121 | 878 | 577 |  |  |  |  |
| C state survival based on Wijngaarde et al., 2020 [55] |  |  |  |  |  |  |  |
| NBS | £162,528,089 | 1,365 | 1,152 | −£108,623,888 | 211 | 402 | −£270,023<br>(dominant) |
| No NBS | £271,151,977 | 1,154 | 749 |  |  |  |  |
| Societal perspective |  |  |  |  |  |  |  |
| NBS | £166,772,287 | 1,346 | 1,140 | −£78,876,005 | 423 | 529 | −£149,074<br>(dominant) |
| No NBS | £245,648,292 | 924 | 611 |  |  |  |  |

*ICER* incremental cost-effectiveness ratio; *LY* life-year; *NBS* newborn screening; *QALY* quality-adjusted life year.

## DISCUSSION

In this economic analysis, NBS for SMA for a cohort of 585,195 newborns identified during a single year and followed over their lifetime was associated with a gain of 529 QALYs and savings of £62,191,531 when compared with no NBS in England. This demonstrates that NBS is dominant (less costly and more effective) compared with no NBS. NBS for SMA would be cost effective and cost saving compared with no NBS for patients with SMA from the perspective of the NHS.

Infants at risk for SMA identified by NBS achieved more motor milestones, improved lifetime health outcomes, and reduced health care costs compared with patients who were clinically diagnosed after symptom onset; therefore, the costs of NBS on the NHS are fully offset by the cost savings associated with early identification and treatment of infants at risk for SMA.

The DSA and PSA demonstrated the robustness of the model and validated the cost-effectiveness outcomes, indicating that NBS for SMA is cost saving for all variations in the sensitivity and scenario analyses. In addition, this model was built on the same Markov structure and applied key assumptions used in the model assessing the cost effectiveness of onasemnogene abeparvovec for SMA type 1, which has been accepted by NICE [37], and other published SMA models [33, 38, 60, 61].

This analysis was performed based on the estimated live births in England; however, including other nations of the United Kingdom, such as Wales, Scotland, and Northern Ireland, would not affect the overall result that NBS for SMA would be a cost-saving health care utilization for the respective NHS. Cost-effectiveness models have been conducted in several other countries, including from the Dutch payer perspective, and have also demonstrated the cost effectiveness of NBS for SMA [33]. Adoption of NBS in the European Union [29] and in the rest of the world [30] is rapidly expanding. For example, approximately 85% of newborns are screened for SMA in the United States [62]. The findings of this economic analysis provide a strong rationale for the introduction of NBS for SMA in England.

Data limitations of the study were mitigated by extrapolating efficacy and survival data. Applying parametric survival extrapolation to estimate long-term patient survival carries a high degree of uncertainty. To verify the survival curves, expert opinion was applied, and the most conservative survival parameters were chosen for the base-case results.

Efficacy was extrapolated from presymptomatic studies, which included patients who were identified via NBS or clinical diagnosis after symptom onset (no NBS). Considering this, we have estimated that approximately 40% of patients with two copies of *SMN2* would be symptomatic at the time of treatment initiation.

The model does not take into consideration the diagnostic journey of patients following symptom onset. A recent Italian study has demonstrated a delay between first symptoms and diagnosis of SMA of 1.94, 5.28, and 16.8 months for patients with SMA types 1, 2, and 3 respectively. This journey included several medical consultations and other examinations such as magnetic resonance imaging, electromyography, or muscle biopsy. Considering these potential additional costs of diagnosis would result in an even more favorable scenario for NBS [63].

In the model, patients with four copies of *SMN2* were considered equal to patients with three copies of *SMN2* in terms of costs and outcomes, because data for patients with four *SMN2* copies treated at birth are critically lacking and no comparison exists for patients treated after symptom onset versus untreated patients. Therefore, we adopted a conservative approach and considered these patients similar to patients with three copies of *SMN2*.

Although the published consensus is to treat patients with four copies of *SMN2*, several countries have adopted a “watch and wait” strategy for these patients. A recent study demonstrated that the economic benefits of NBS for patients with four copies of *SMN2* were substantially less than for patients with three copies [28]. From a clinical perspective, however, recent data from Germany demonstrated that five of seven patients with four copies of *SMN2* may develop irreversible symptoms of SMA before the age of 4 years [64]. It must be noted that *SMN2* copy number quantification is not entirely standardized and that significant inter- [65] or intra- [66] laboratory differences may be observed, especially for the higher copy numbers. Nevertheless, it is very unlikely that this would significantly alter the conclusion of this model or other health economic assessments because different studies converge to demonstrate a similar percentage of copy numbers in the SMA subpopulation.

More research is needed to identify long-term costs for surviving patients, specifically the costs associated with the walking and sitting health states. In the model, for all available treatment options, the same costs were applied per health state to avoid bias towards any of the treatments. The sensitivity of final economic outcomes to the magnitude of resource use costs by health states was evaluated in deterministic sensitivity analysis (*see* **Fig. 2**), which indicated in all cases NBS as a dominant option over clinical diagnosis (no NBS).

## CONCLUSIONS

NBS for SMA in England is less costly and more effective than a strategy without NBS. Therefore, our findings support the inclusion of NBS for SMA in England.

## Data Availability

All data generated or analyzed during this study are included in this article [and its supplementary information files].

## DECLARATIONS

### Funding

This study was funded by Novartis Gene Therapies, Inc.

### Author Contributions

Conceptualization: MB, DW, IK Methodology: MB, DW Screening/data extraction: MB, DW Writing: The first draft was prepared by DW and RH and reviewed by all authors. Review: All authors have read and approved the final manuscript.

### Disclosures

**DW** and **RH** are full-time employees of Clarivate and consultants for Novartis Gene Therapies, Inc. **LS** has received personal compensation for consultancy for Biogen, Biophytis, Cytokinetics, Dynacure, Novartis Gene Therapies, Inc., Novartis, Roche, Santhera, Sarepta Therapeutics, and Zentech. He has received research support from Novartis Gene Therapies, Inc., Biogen, Dynacure, and Roche; and he leads the Belgium and UK NBS funded by Biogen, Novartis, and Roche. **IK** and **MB** are employees of Novartis Gene Therapies and own stock/other equities.

### Compliance with Ethics Guidelines

This article is based on a cost-utility analysis using a combination of decision tree and Markov model structures, and does not contain any new studies with human participants or animals performed by any of the authors.

### Data Availability

All data generated or analyzed during this study are included in this published article/as supplementary information files.

### Consent to Participate

N/A

### Prior Presentation

This data has been previously presented as a poster at ISPOR’s 25^th^ annual European congress, held on November 6–9, 2022, Vienna, Austria.

## Notes

### Summary of Updates

Peer review comments have been addressed, including an update to the Discussion to clarify the source of live births and the authors' methods for including four copy patients. In addition, the title has been changed to remove Wales as this analysis and model focused on England only. This adjustment has been carried through the entire paper.

