## Supplemental material for "Cost Effectiveness of Newborn Screening for Spinal Muscular Atrophy in England"

[1]Health Economics, Clarivate, Frankfurt am Main, Germany; [2]MDUK Oxford Neuromuscular Centre & NIHR Oxford Biomedical Research Centre, University of Oxford, Oxford, UK; [3]Neuromuscular Center of Liège, Department of Paediatrics, Hospital and University of Liège, Liège, Belgium; [4]Novartis Gene Therapies, London, UK; [5]Health Technology Assessment, Clarivate, Manchester, UK; [6]Novartis Gene Therapies GmbH, Rotkreuz, Switzerland

**Corresponding Author:**

Diana Weidlich, MSc

Clarivate

Mayfarthstrasse 11

Frankfurt am Main 60314

Germany

### A. Cost inputs for running scenario analysis: societal perspective

**Table S1. Societal costs: cost summary per health state**

| Costs (£) | Health state E | Health state D | Health state C | Health state B | Health state A |
|---|---|---|---|---|---|
| Annual indirect costs | 22,999 | 22,999 | 16,560 | 0 | 0 |
| Annual carer costs | 48,004 | 48,004 | 30,243 | 0 | 0 |
| Onasemnogene abeparvovec /Risdiplam - transport costs for treatment | 21 | 16 | 10 | 5 | 5 |
| Nusinersen - transport costs for treatment (plus dosing related visits calculated below) | 21 | 16 | 16 | 16 | 16 |
| BSC - transport costs for treatment | 31 | 31 | 31 | 0 | 0 |

*BSC* best supportive care.

**Table S2. Societal costs: indirect cost inputs**

| Indirect costs | Input value |
|---|---|
| Average labor force participation rate (%) [1] | 75 |
| Average working hours per year [2] | 1,934 |
| Percentage of work hours lost per health state | |
| Health state E [3] | 100 |
| Health state D [3] | 100 |
| Health state C [3] | 72 |
| Health state B[a] | 0 |
| Health state A[a] | 0 |
| Average hourly wage (£) [2] | 16 |
| Productivity cost per hour (£) [4] | 34 |
| Start working age (years)[a] | 18[a] |
| End working age (years) [5] | 68 |

[a]Based on assumption.

**Table S3. Societal costs: caregiver cost inputs**

| Caregiver costs | Input value |
|---|---|
| Average hourly wage for a caregiver (£) [6] | 33 |
| Percentage of patients requiring a carer per health state[a] | |
| Health state E | 100 |
| Health state D | 100 |
| Health state C | 63 |
| Health state B | 0 |
| Health state A | 0 |

[a]Based on assumption.

**Table S4. Societal costs: transport cost inputs**

| | Input value |
|---|---|
| Percentage of patients that incur transport costs[a] | 100 |
| Average distance to academic hospital (kilometer) [7] | 9.0 |
| Cost per kilometer (£) | 0.3 |
| Parking per visit (£) [8] | 0.0 |
| Cost per kilometer taxi (£) [9] | 4.9 |
| Total transport costs per visit (£) [10] | 5.2 |

[a]Based on assumption.

**Table S5. Societal costs: average number of hospital visits per year**

| | Average number of visits (per year) onasemnogene abeparvovec /risdiplam[a,b] | Average number of visits (per year) nusinersen (plus dosing visits)[a,b] | Average number of visits (per year) BSC[a] |
|---|---|---|---|
| Health state E | 4 | 4 | 6 |
| Health state D | 3 | 3 | 6 |
| Health state C | 2 | 3 | 6 |
| Health state B | 1 | 3 | 0 |
| Health state A | 1 | 3 | 0 |

*BSC* best supportive care.

[a]Based on assumption.

[b]Based expert opinion.

### B. Results

#### B.1. Proportion of SMA patients, alive and by level of motor milestone achievement over time

To provide additional insight on the main drivers of health gains and health care cost savings associated with NBS and early treatment, the percentage of patients residing in each of the six health states of the model was computed at different time points to follow the children's development over time. The results of this analysis under NBS and no NBS from 5 years to 30 years of follow-up are provided in **Figs. S1** and **S2** below.

The main key findings of the analysis are the following:

- With NBS and early treatment, from the age of 5 years old onwards, approximately 80% of children with SMA will sit and walk independently, as opposed to approximately 20% of children with no NBS who are clinically diagnosed.
- In the current treatment situation with no NBS in place, for most patients with SMA, sitting will be the highest motor milestone that they will ever achieve.
- Without NBS and early treatment, less than 50% of children with SMA are expected to live beyond the age of 30, compared with 80% of patients with NBS and early treatment. In fact, with NBS and early treatment, 70% of patients are expected to reach the age of 70 versus less than 20% of patients diagnosed symptomatically.

**Figure S1. Percentage of SMA patients, alive and by motor milestone achievement over time, with NBS and early treatment**

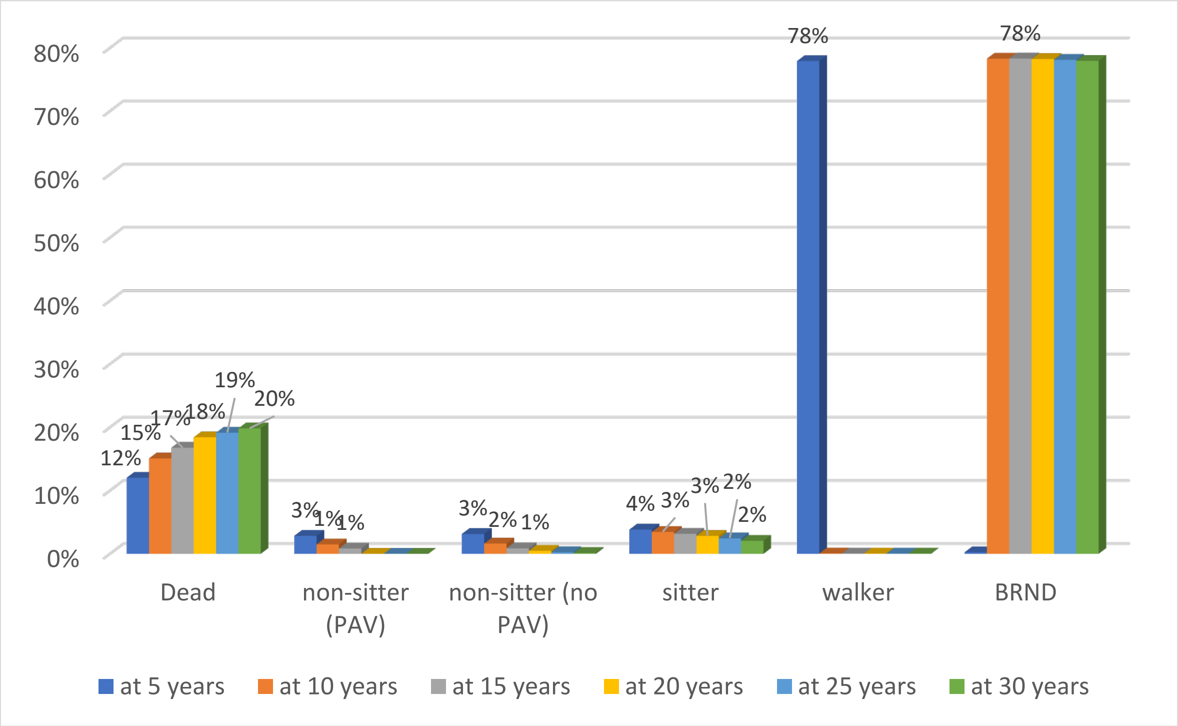


*BRND* broad range of normal development; *NBS* newborn screening; *PAV* permanent assisted ventilation.

**Figure S2. Percentage of SMA patients, alive and by motor milestone achievement over time, in the current situation: no NBS and treatment at symptom onset**

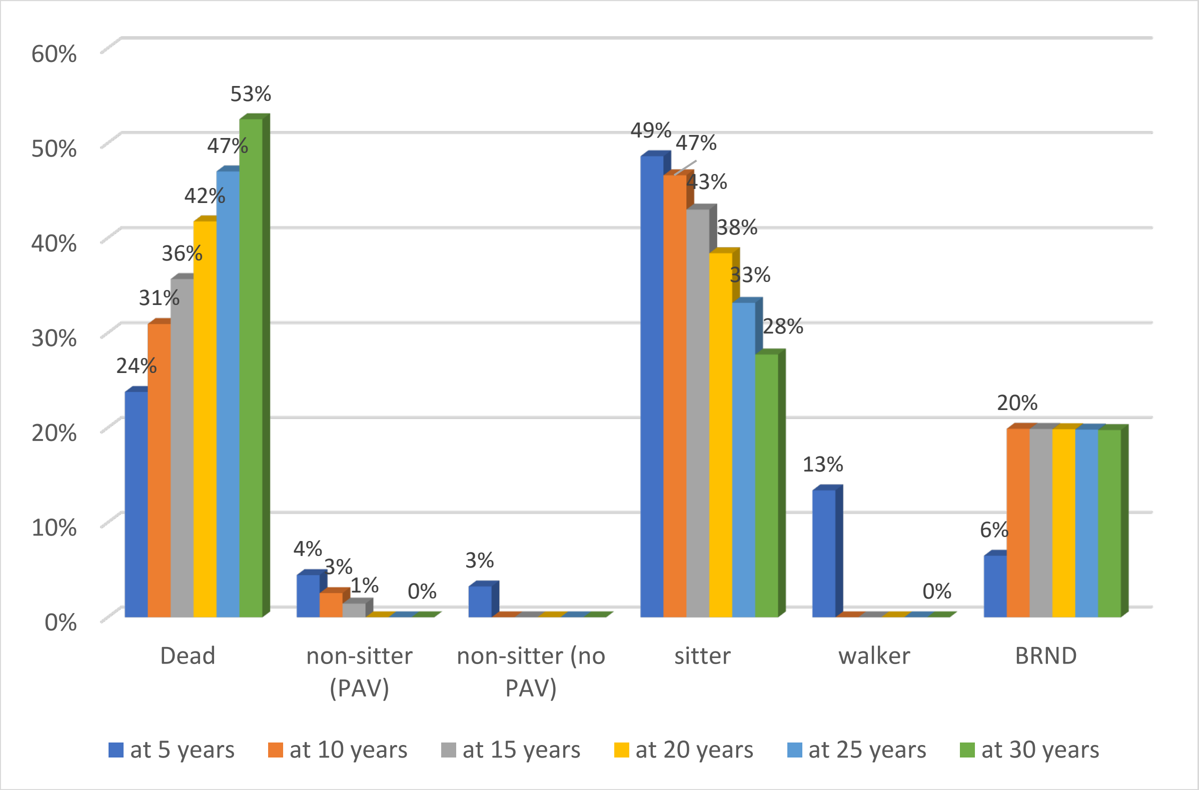


*BRND* broad range of normal development; *NBS* newborn screening; *PAV* permanent assisted ventilation.

**Table S6. Percentage of SMA patients at age 70, alive and by motor milestone achievement over time, under NBS and early treatment compared with no NBS and treatment at symptom onset**

| | Dead | Non-sitter (PAV) | Non-sitter (no PAV) | Sitter | Walker | BRND | Sum |
|---|---|---|---|---|---|---|---|
| **SMA patients identified via NBS and treated immediately upon diagnosis** | **33%** | 0% | 0% | 0% | 0% | 67% | 100% |
| **SMA patients treated at symptom onset** | **82%** | 0% | 0% | 2% | 0% | 16% | 100% |

*BRND* broad range of normal development; *NBS* newborn screening; *PAV* permanent assisted ventilation; *SMA* spinal muscular atrophy.

### B.2. Economic outcomes per SMA patient

**Table S7. Undiscounted and discounted disaggregated costs and QALYs per SMA patient under NBS and no NBS**

| | Costs per SMA patient | | |
|---|---|---|---|
| | **NBS** | **No NBS** | **Increment** |
| **Disaggregated cost results — undiscounted** | | | |
| Screening costs[a] | £46,200 | £1,200 | £45,000 |
| Drug acquisition and administration costs | £4,668,496 | £6,132,232 | –£1,463,736 |
| Medical care costs | £428,274 | £1,396,914 | –£968,640 |
| Total undiscounted cost saving net of screening costs | | | –£2,387,376 |
| **Disaggregated cost results — discounted at 3.5% p.a.** | | | |
| Screening costs[a] | £46,200 | £1,200 | £45,000 |
| Drug acquisition and administration costs | £2,346,391 | £2,949,844 | –£603,452 |
| Medical costs | £342,703 | £847,000 | –£504,297 |
| **Total discounted cost saving net of screening costs** | | | **–£1,062,749** |
| | **QALYs per SMA patient** | | |
| | **NBS** | **No NBS** | **Increment** |
| **Total undiscounted QALY gain** | 56 | 24 | **32** |
| Total QALY gain discounted at 3.5% p.a. | 19 | 10 | 9 |

*NBS* newborn screening; *p.a* per annum; *QALY* quality-adjusted life year; *SMA* spinal muscular atrophy.

[a]Screening costs include £4.54 for the heel-prick test and £1,200 for the confirmatory genetic test. A total of 58.8 SMA patients receive the confirmatory genetic test under both NBS and no NBS. Under NBS, 585,195 newborns receive the heel-prick test.

Newborn screening and early treatment are expected to provide infants at risk for SMA an additional 32 (undiscounted) years at full health when compared with the current situation in which patients with SMA are treated at symptom onset.

Implementing NBS and providing early treatment are also expected to generate an overall discounted cost savings, net of the cost of screening all newborns, of more than £1 million per SMA patient. These savings capture savings in medical care costs, such as care during hospital admissions, breathing equipment, and other costly health care services, and a reduction in drug acquisition costs because of different treatment patterns used for treating presymptomatic versus symptomatic patients (see **Table 1** in main document).
